# National trends in rates of undiagnosed HIV in key populations in Australia: a retrospective observational analysis from 2008 to 2019

**DOI:** 10.1101/2022.09.23.22280259

**Authors:** Richard T. Gray, Hawa Camara, Laila Khawar, Andrew Grulich, Rebecca Guy, Skye McGregor, Nicholas Medland

**Affiliations:** The Kirby Institute, UNSW Sydney, Sydney, Australia

**Keywords:** HIV undiagnosed, total diagnosed fraction, yearly diagnosed fraction, case detection rate

## Abstract

**Introduction:** Determining the proportion of people living with HIV who are undiagnosed is critical for Australia to accurately assess the country’s progress toward UNAIDS’ 95-95-95 interim targets by 2025 and progress with elimination goals. We aimed to investigate the utility of two additional measures, the Yearly Diagnosed Fraction, and the Case Detection Rate, in elucidating trends over time in the rate of undiagnosed HIV.

**Methods:** Using routinely collected national HIV surveillance data, we produced estimates for the number living with undiagnosed HIV and the number of new HIV infections using the European Centre for Disease Protection and Control (ECDC) HIV modelling tool to calculate the Total Diagnosed Fraction (TDF), the Yearly Diagnosed Fraction (YDF), and the Case Detection Rate (CDR) from 2008 to 2019 for Australian-born and overseas-born individuals who have reported having male-to-male sexual contact, and heterosexual women and men. We calculated rate ratios using Poisson Regression models to compare trends for each sub-population.

**Results:** Over 2008-2019, each metric for the Australian-born male-to-male sexual contact group improved consistently over the period with the case detection rate rising above one in 2013. The total diagnosed fraction for the overseas born group fell slightly from 85.0% to 81.9%, the yearly diagnosed fraction fell from 23.1% to 17.8% and the case detection rate stayed below one, falling from 0.74 to 0.63. In the heterosexual group, women and men had consistent increasing trends for the total diagnosed fraction and yearly diagnosed fraction but with women having consistently higher estimates than men, 92.6% vs 80.8% and 26.3% vs 17.4%, respectively in 2019. However, heterosexual men had a declining case detection rate, falling below one in 2019 (0.83), compared to an increase for women.

**Conclusion:** The additional metrics presented provide important information on Australia’s progress toward HIV elimination. The more dynamic changes in the undiagnosed population seen highlight diverging trends for key populations with growing undiagnosed populations not seen in the total diagnosed fraction. Periodic trend analyses will help strengthen the use of the metrics and ease of interpretation for national surveillance reporting.

## Introduction

The HIV diagnosis and care cascade is used internationally to demonstrate progress with key HIV care elements, reflected in UNAIDS 95-95-95 targets to end the AIDS epidemic by 2030 and to which Australia has committed.[1,2] The first step in the HIV cascade, and UNAIDS target, is the proportion of people living with HIV who have been diagnosed, otherwise known as the total diagnosed fraction, with the goal of achieving the target of 95% by 2025.[3] Undiagnosed HIV contributes to more than half of new HIV infections globally each year.[4] People with undiagnosed HIV are at greater risk of ill health and onward transmission.[5] To reduce HIV incidence and morbidity, it is essential to ensure timely diagnosis of people living with undiagnosed HIV and initiate effective and lifesaving antiretroviral therapy (ART) to ensure they achieve viral suppression, effectively reducing the risk of HIV transmission to HIV-negative sexual partners to zero.[1,4]

The first step of the HIV cascade is calculated using a denominator that is the estimated total number of people living with HIV, which includes those not yet diagnosed as well as those on treatment and who have achieved viral suppression, the second two 95% UNAIDS targets. For established HIV epidemics where most individuals living with HIV have already been diagnosed and linked to treatment, any changes in the number undiagnosed (for example due to an increase in incidence or HIV testing) will have a limited relative impact on the numerator and denominator in the total diagnosed fraction calculation and hence will be obscured. Measures which more directly capture the dynamics of the undiagnosed population may better reflect the public health goal of getting people diagnosed and linked to care, and may provide additional information for monitoring progress toward HIV elimination goals.[6–8]

The Australian HIV Cascade shows that at the end of 2019, there were an estimated 28,680 people living with HIV, of which 25,880 had been diagnosed (90.2%) and 2,800 people (9.8%) were living with undiagnosed HIV.[9] The total diagnosed fraction has only increased slightly in the past 5 years despite increases in testing, early diagnosis and treatment and the scale-up of HIV pre-exposure prophylaxis (PrEP). However, the relatively large denominator of people living with HIV may have obscured the relatively smaller changes in the number undiagnosed making up the numerator. Obscured, also are large variations in sub-populations such as overseas born gay, bisexual, and other men who have sex with men.

The Australian HIV cascade estimates for the total diagnosed fraction are obtained annually using HIV surveillance data and the European Centre for Disease Protection and Control (ECDC) HIV Modelling Tool.[10,11] This tool provides further outputs, such as the number of new infections and the estimated time between infection and diagnosis, that could be used to generate additional indicators, complementing the total diagnosed fraction, to track changes in the undiagnosed population. Two of these potential indicators are the yearly diagnosed fraction and the case detection rate which do not include the large number of diagnosed individuals in the numerator or denominator. The yearly diagnosed fraction is the fraction of all people who need to be diagnosed, who have been diagnosed, during a year. The case detection rate is the ratio of diagnoses to new infections and indicates whether the number of people with undiagnosed HIV increases or decreases. In this retrospective study we used the same HIV cascade data sources to calculate these two indicators to see if they better capture trends and variations across populations in the undiagnosed population.

## Methods

For our study, we used national surveillance data from the years 2008 to 2019 (prior to the COVID-19 pandemic).

### Study populations

Routinely collected HIV notification surveillance data including, gender, country of birth, and likely mode of HIV exposure were used to define the sub-populations of interest—people reporting male-to-male sexual contact as HIV exposure risk, stratified by overseas born and Australian born, and people reporting heterosexual contact as exposure risk, stratified by gender (male and female). We selected these sub-populations because notifications reporting male-to-male sexual contact account for the highest proportion of notifications in Australia, and recent epidemiological data have indicated a divergence in rates between people born in Australian versus overseas [9]; and because notifications reporting heterosexual contact have not experienced the same decline as seen among individuals reporting male-to-male sexual contact and have fluctuated between 19-25% of the total notifications during the 2008-2019 period.

### Defining indicators

We calculated estimates for the following indicators for each sub-population as defined below.

#### 1. Total Diagnosed Fraction (TDF)

The total diagnosed fraction is a widely used HIV cascade measure and refers to the proportion of all people living with HIV (PLHIV) who have been diagnosed and is calculated for each year as follows:

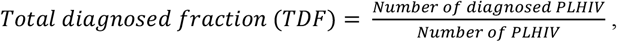

where the numerator and denominator are the cumulative number at the end of the year and PLHIV includes all people living with HIV (both diagnosed and undiagnosed).

#### 2. Yearly Diagnosed Fraction (YDF)

The yearly diagnosed fraction is an annual measure of the proportion of all of those living with undiagnosed HIV who receive a diagnosis that year.[6,8] The inverse of the YDF reflects the number of years it would take to diagnose all undiagnosed people if there were no new infections. For example, a yearly diagnosed fraction of 0.2 implies it would take 5 years to diagnose all people living with HIV in that population. Ideally, the YDF should be as close to one as possible. Should every person living with undiagnosed HIV be diagnosed within a year, the YDF would equal one or equivalently 100% when converted to a percentage. A low YDF also indicates that a higher proportion of individuals were diagnosed with late HIV, which would not be apparent from the cascade. The YDF is calculated for each year as follows:

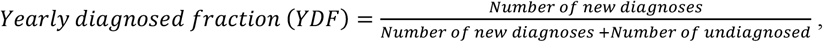

where the number of new diagnoses is the number during the year and number undiagnosed is the total number undiagnosed at the end of the year. We multiplied the YDF calculated with this equation by 100 to obtain a percentage.

#### 3. Case Detection Rate (CDR)

The case detection rate is a key indicator in tuberculosis research [12,13]. It is the ratio of new infections to new diagnoses during each year. If the number of new infections equals the number of new diagnoses, the CDR is equal to one and the number of people who have not been diagnosed is unchanged. If the CDR is greater than one, then the number of undiagnosed individuals will decrease. The CDR for each year is calculated as:

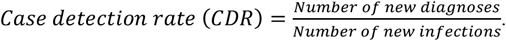

Ideally, the CDR should be above one and as high as possible to reduce the population living with undiagnosed HIV as quickly as possible.

### Estimating new HIV diagnoses, number undiagnosed, number diagnosed, and new infections

We used the ECDC HIV modelling tool to estimate the number of people living with undiagnosed HIV, the number of new HIV infections each year, and the total number of people living with HIV for the TDF, YDF, and CDR calculations. This tool is a multi-state back-calculation model that fits diagnoses over time using data on new HIV and AIDS diagnoses, estimates for the number of annual deaths and emigrations within people living with diagnosed HIV, and estimates for the rate of CD4+ T-cell decline.[10,11] The tool is used for national surveillance reporting in Australia and produces several outputs including the proportion of a population living with undiagnosed HIV and the annual number of new HIV infections.[9] A detailed description of how the ECDC tool is used for HIV surveillance in Australia is provided elsewhere.[9,14]

The ECDC tool requires inputs of annual estimates for the number of HIV and AIDS diagnoses overall and by CD4 count category (>= 500, 350-499, 200-349, < 200 cells/mm^3^), and the number of all cause deaths and emigrations from Australia among people living with diagnosed HIV. We estimated the annual number of HIV diagnoses among each sub-population using routinely collected national HIV surveillance data (including date of HIV diagnosis, CD4+ T-cell count at diagnosis, date AIDS diagnosis, gender, country of birth, year of arrival in Australia, and HIV exposure category) and statistical imputation to fill in missing data (with averages from 10 imputed data sets produced used for our results) and adjusting for duplicates using a previously developed algorithm.[9,15] For reporting purposes, we rounded the resulting estimates to the nearest whole number. We used all-cause death rates estimated from a linkage study between national HIV and death registers until 2003, and a large national cohort study of people living with HIV since.[2,16]

The number of diagnosed people who leave Australia each year was estimated using two emigration adjustments. First, data from six-month follow up of all people notified with HIV in NSW, Australia’s most populous state, was used to estimate short-term emigration.[9] Second, longer term emigration, was estimated using migration data from the Australian Bureau of Statistics.[17] We assumed that long term emigration rates for people living with HIV were the same as for the general population. The annual number of people living with diagnosed HIV is then obtained by adding annual diagnoses and subtracting estimated deaths and emigrations which are obtained by applying the death rates and emigration adjustments to the previous year’s estimate for the number living with diagnosed HIV. The resulting annual diagnoses, all-cause deaths, and overseas migrations were then inputted into the ECDC tool. Estimates and uncertainty ranges for our analyse were generated from 100 runs of version 1.3.0 of the ECDC HIV Modelling Tool. The number of people with undiagnosed HIV was obtained by applying the proportion undiagnosed to the estimated number living with diagnosed HIV at the end of each year.

Some people living with HIV in Australia will have previously received an HIV diagnosis in another country. These people only enter the national HIV registry and become officially notified when they receive a confirmatory diagnosis in Australia. Confirmation of HIV status is routinely performed at point of entry into clinical care and the notification record includes information on previous diagnosis overseas—which is primarily obtained through self-report.[9] We assumed people living with HIV in Australia previously diagnosed overseas are aware of their HIV status and hence part of the diagnosed population (even if they are not in care or have not been notified within Australia). To estimate annual new infections and the proportion diagnosed we used the ECDC tool with previously diagnosed overseas excluded from diagnoses. Note this approach could still overestimate new infections and the number undiagnosed if a substantial proportion of people who acquired HIV overseas are first diagnosed in Australia and categorised as acquiring HIV in Australia.[18] For these estimates we produced a 95% CI using 100 bootstrapped fits produced by the ECDC tool.

### Statistical Analysis

To assess time trends in TDF, YDF and CDR across key populations, we used Poisson regression models to calculate average annual rates of change (average annual trend), presented as rate ratios (RR), over time, between 2008 and 2019. For example, an RR of 0.80 signified a 20% average annual decrease. Difference at baseline (baseline rate ratio) and differences in rate of change (ratio of difference or summary rate ratio - SRR) by gender (heterosexual population) and by place of birth (male-to-male sexual contact) were quantified in these models. This was done by including interaction terms between calendar year and gender/place of birth. The Poisson regression models were calculated using the STATA Statistical Software (version 16.0; college station, TX: StataCorp LLC).

No ethical approval was required for this study because we used modelling estimates generated through routine surveillance mechanisms. The code used to produce the numerator and denominator estimates for each indicator and the inputs for the ECDC HIV modelling tool is available online and uses R version 4.2.1. [19,20] Legislation prohibits sharing of HIV notifications data, however, aggregate data and results are available in the online repository.[20]

## Results

Between 2008 and 2019, there were 14,296 HIV notifications reported, of which an estimated 11,240 were people who were first diagnosed with HIV in Australia. Of these people, 5,543 (49%) were Australian-born individuals who had reported having male-to-male sexual contact, 3,023 (27%) were men with the same exposure category born overseas. Amongst the 2,673 people reporting heterosexual contact, there were 1,122 women (10%) and 1,551 (14%) men. See Table 1 for further detail.

**Table 1.**
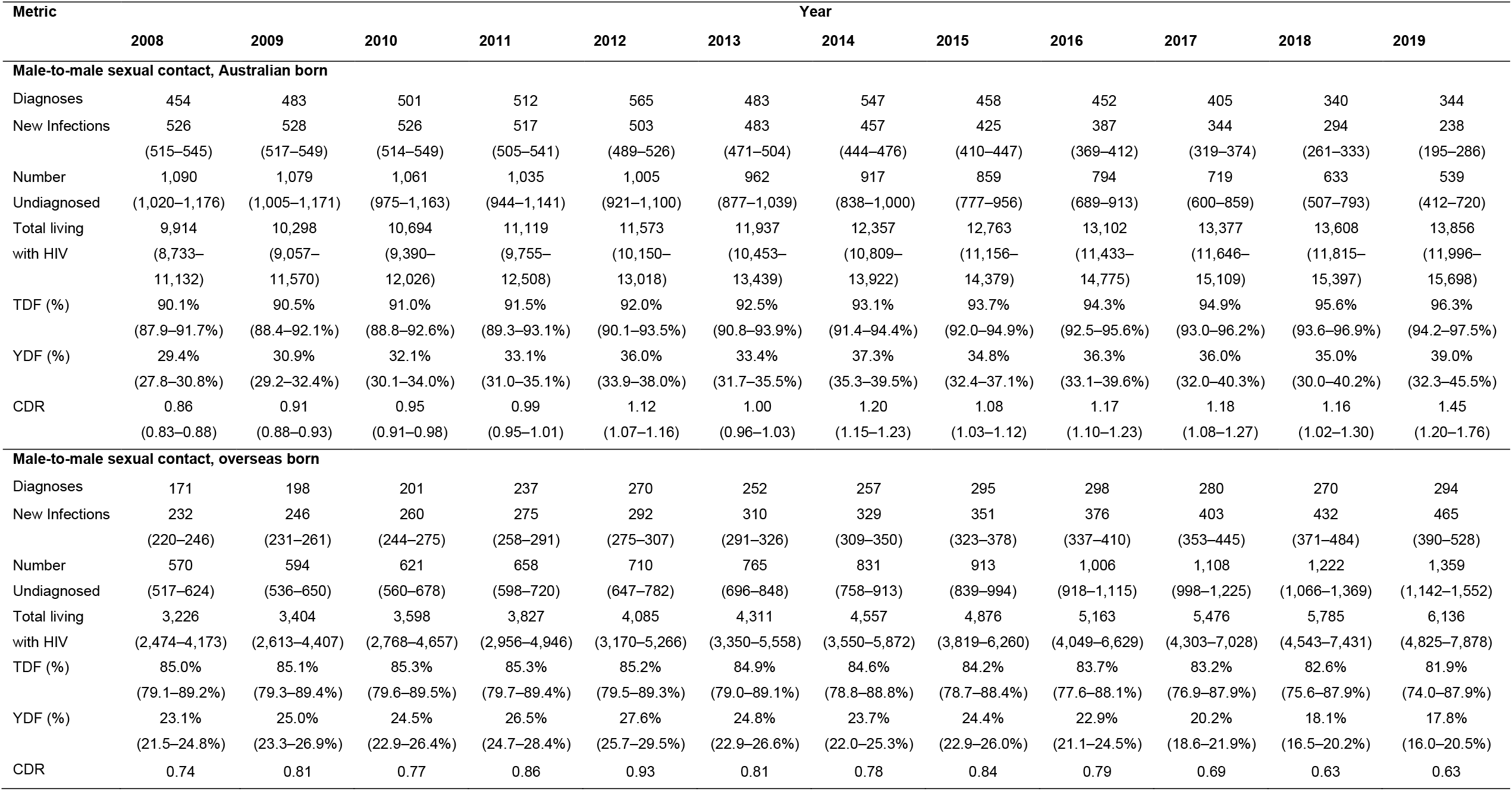
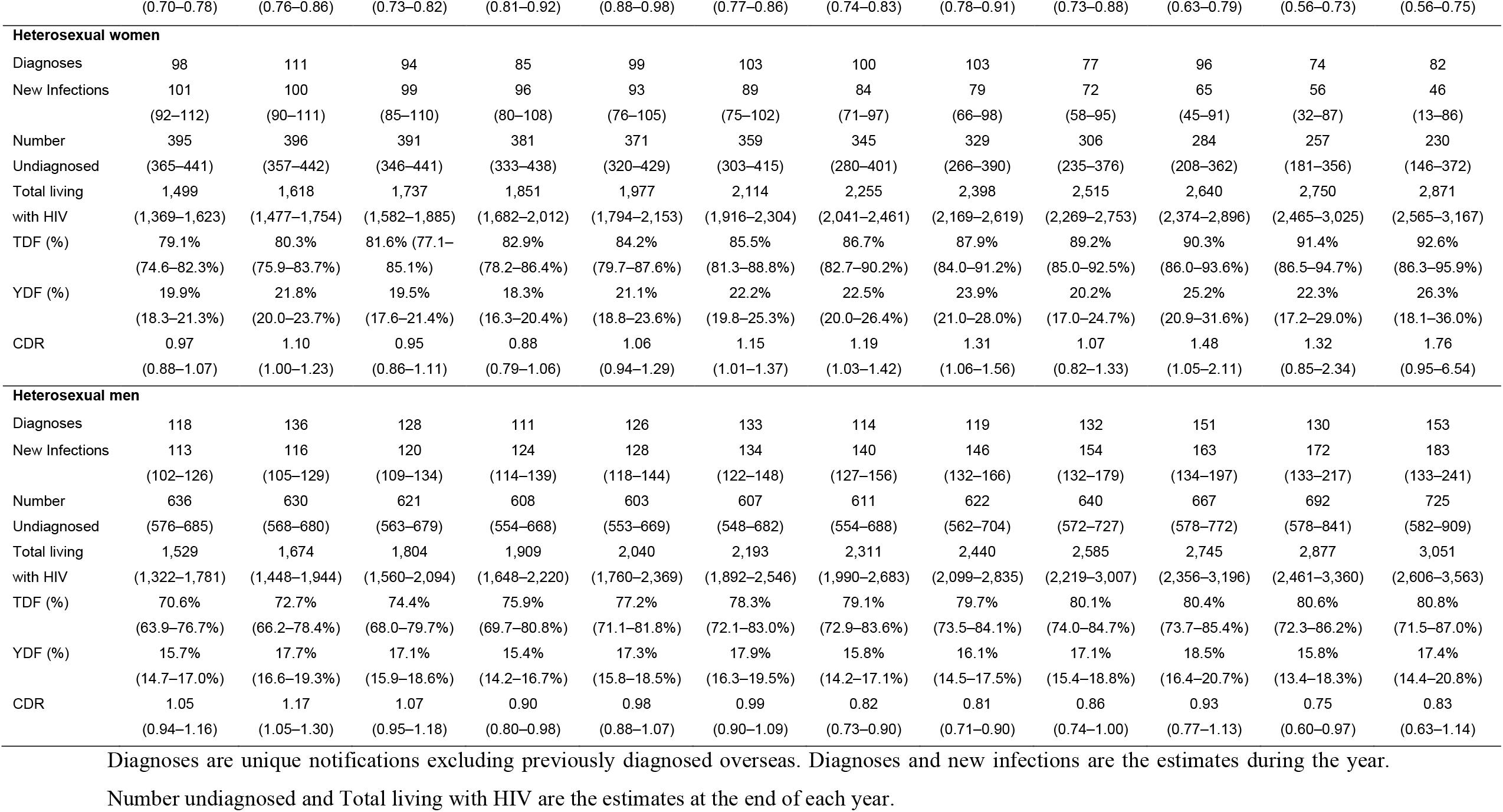
Annual estimates and ranges for each sub-population over 2008 - 2019.

### Male-to-male sexual contact

The number of diagnoses among Australian-born men reporting male-to-male sexual contact increased from 454 in 2008 to 565 in 2012 and then decreased to 344 in 2019, while the number of diagnoses who were overseas-born increased steadily from 171 in 2008 to 295 in 2015 then remained stable with 294 diagnoses in 2019. Over the same period, the estimated number of undiagnosed Australian-born men with this exposure risk decreased from 1,090 to 539. In the overseas born the number undiagnosed increased from 570 to 1,359. The estimated number of new infections decreased from 526 to 238 and increased from 232 to 465 in the Australian born and overseas born, respectively. See Table 1.

Among Australian-born men who reported having male-to-male sexual contact, the proportion living with HIV who had been diagnosed (TDF) increased from 90.1% to 96.3% (average annual rate ratio of RR:1.01 95% CI 1.00-1.01, p<0.0001. The YDF, the proportion of those undiagnosed who received a diagnosis in a year, increased from 29.4% to 39.0% (with a steady increase from 2008 to 2014, then a plateau before a large rise in 2019) with an average annual increase of 2% (RR: 1.02, 95% CI 1.01-1.03, p<0.0001). This means, inversely, it would take more than two and a half years to diagnose all undiagnosed people if there were no new infections. The CDR (the ratio of new diagnoses to new infections during a year) rose from 0.86 to 1.45 (4% average annual increase, RR: 1.04, 95% CI 1.03-1.04, p<0.0001). The 2019 estimate of 1.45 indicates that for every two people who acquired HIV that year approximately three undiagnosed individuals were tested and diagnosed.

In those born overseas, the TDF fell slightly from 85.0% at 81.9% (RR: 0.996 95%, CI 0.994-0.999, p=0.0025; Figure 1), the YDF initially increased from 23.1% to 27.6% in 2012 before falling to 17.8% in 2019 (RR: 0.97, 95% CI 0.96-0.98, p<0.0001) meaning it would take more than 5 years to diagnose all undiagnosed people if there were no new infections. The CDR followed a similar trend rising from 0.74 in 2008 to 0.93 in 2012 and then declining to 0.63 in 2019, being lower than the estimates for Australian-born men throughout (average annual increase 1%, RR: 0.98, 95% CI 0.97-0.99, p<0.0001) (see Figure 1). Having a CDR less than one during the whole period means the number undiagnosed has been continually increasing. The biggest differences in the trends between the Australian-born and overseas-born men were in the YDF (SRR: 1.05, 95% CI 1.04-1.07, p<0.0001) and CDR (SRR: 1.06, 95% CI 1.05-1.07, p<0.0001) with only a slight (but statistically significant) difference in trends for the TDF (SRR: 1.01, 95% CI 1.00-1.01, p<0.0001). See Table 2.

**Figure 1:**
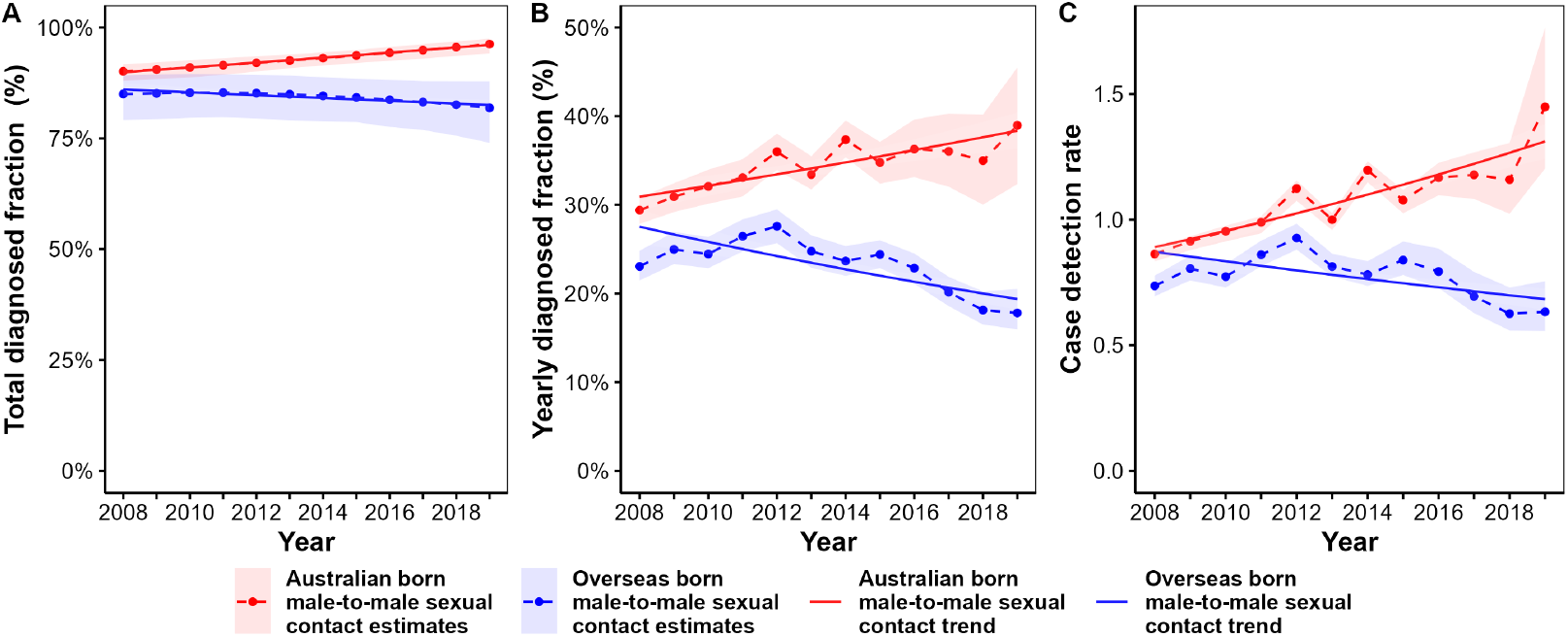
(A) Total Diagnosed Fraction, (B) Yearly Diagnosed Fraction, and (C) Case Detection Rate for individuals having reported male-to-male sexual contact, Australian-born versus Overseas-born.

**Table 2.** Trends in Total Diagnosed Fraction (TDF), Yearly Diagnosed Fraction (YDF), and Case Detection Rate (CDR) for each sub-population over 2008 - 2019.

|  | **TDF** |  | **YDF** |  | **CDR** |  |
| --- | --- | --- | --- | --- | --- | --- |
|  | Average annual trend | Ratio of difference | Average annual trend | Ratio of difference | Average annual trend | Ratio of difference |
|  | RR (95% CI)<br>p value | SRR (95% CI)<br>p value | RR (95% CI)<br>p value | SRR (95% CI)<br>p value | RR (95% CI)<br>p value | SRR (95% CI)<br>p value |
| **Male-to-male sexual contact** |  |  |  |  |  |  |
| Australian born | 1.01 (1.00-1.01)<br>p<0.0001 | 1.01 (1.00-1.01)<br>p<0.0001 | 1.02 (1.01-1.03)<br>p<0.0001 | 1.05 (1.04-1.07)<br>p<0.0001 | 1.04 (1.03-1.04)<br>p<0.0001 | 1.06 (1.05-1.07)<br>p<0.0001 |
| Overseas born | 0.996 (0.994-0.999)<br>p=0.0025 | Reference | 0.97 (0.96-0.98)<br>p<0.0001 | Reference | 0.98 (0.97-0.99)<br>p<0.0001 | Reference |
| **Heterosexuals** |  |  |  |  |  |  |
| Women | 1.01 (1.01-1.02)<br>p<0.0001 | 1.00 (0.99-1.01)<br>p=0.1930 | 1.02 (1.00-1.04)<br>p=0.0200 | 1.02 (0.99-1.04)<br>p=0.1201 | 1.05 (1.03-1.06)<br>p<0.0001 | 1.08 (1.05-1.10)<br>p<0.0001 |
| Men | 1.01 (1.01-1.02)<br>p<0.0001 | Reference | 1.00 (0.99-1.01)<br>p=0.6957 | Reference | 0.97 (0.96-0.99)<br>p=0.0001 | Reference |

CI = Confidence interval; RR = Rate ratio; SRR = Summary rate ratio.

### Heterosexual women and men

The number of heterosexual women first diagnosed with HIV in Australia ranged between 74 and 111 over 2008-2019. The number of diagnosed heterosexual men increased from 118 to 153. Over the same period, the estimated number of people undiagnosed went from 395 to 230 in women and from 636 to 725 in men and the estimated number of new infections from 101 to 46 and from 113 to 183, respectively. See Table 1.

Among heterosexual women the percentage who had been diagnosed (TDF) increased from 79.1% to 92.6% (an average annual increase of 1%, RR: 1.02, 95%, CI 1.01-1.02, p<0.0001). The YDF increased from 19.9% to 26.3% (average annual increase of 4%, RR: 1.02, 95% CI 1.00-1.04, p=0.02) and the CDR increased from 0.97 to 1.76 (average annual increase of 5%, RR = 1.05 95% CI 1.03-1.06, p<0.0001). This means the time to diagnose all HIV-positive women has reduced from five to four years and for every four heterosexual woman who acquired HIV in 2019 seven undiagnosed women were diagnosed. Among heterosexual men, the values of the metrics were lower with the TDF increasing from 70.6% to 80.8% (average annual increases 1%, RR: 1.01, 95% CI 1.01-1.02, p<0.0001), the YDF increasing from 15.7% to 17.4% (RR: 1.00, 95% CI 0.99-1.01, p=0.696), and the CDR decreasing from 1.05 to 0.83 (average annual decrease of 3%, RR:0.97, 95% CI 0.96-0.99, p=0.0001). The value of the CDR in 2019 suggests the number of undiagnosed heterosexual men is growing. The biggest difference in the metric trends between heterosexual women and men was for the CDR (SRR: 1.08, 95% CI 1.05-1.10, p<0.0001) with no difference in trends for the TDF (SRR: 1.00, 95% CI 0.99-1.01, p=0.1930) and only a small difference in the YDF trends for each gender (SRR: 1.02, 95% CI 0.99-1.04, p=0.1201). See Tables 1-2 and Figures 2A-C.

**Figure 2.**
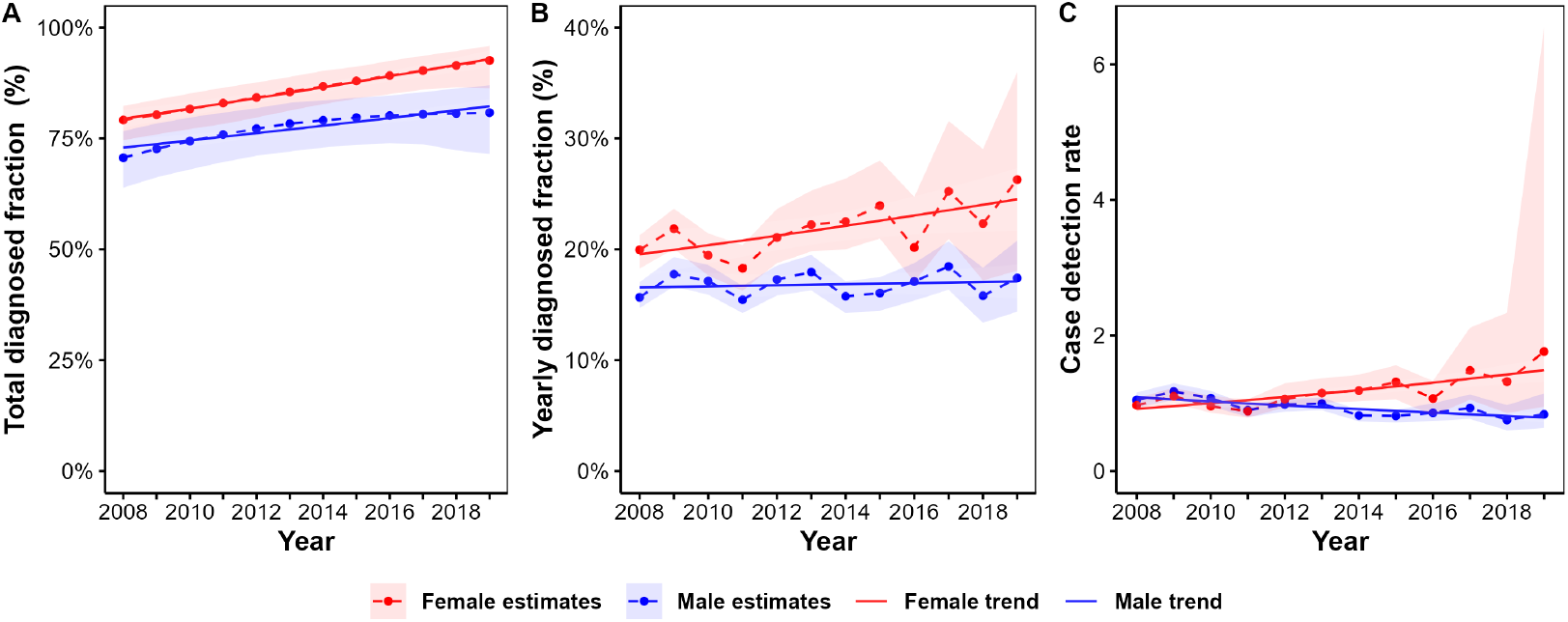
(A) Total Diagnosed Fraction, (B) Yearly Diagnosed Fraction, and (C) Case Detection Rate for Heterosexual women and men.

## Discussion

In comparing trends in three different metrics describing undiagnosed HIV among key sub-populations in Australia we found much larger observed differences in the Yearly Diagnosed Fraction and the Case Detection rate than in the Total Diagnosed Fraction. In men who report male-to-male sexual contact, a slight trend in the diagnosed fraction obscured large changes between Australian born and overseas born in case detection and yearly diagnoses. In heterosexuals, there was no significant trend in the diagnosed fraction despite divergence in case detection.

Among Australian-born men who reported male-to-male sexual contact, the total diagnosed fraction (TDF) increased from 90% to 96% from 2008-2019, the only sub-population who has surpassed the global target of 95%. In the same group, the case detection rate (CDR) increased substantially to approximately 1.45 by 2019 meaning the fall in the number with undiagnosed HIV is accelerating. Even though the yearly diagnosed fraction (YDF) increased to 39% over the study period, this means it would still take over two and a half years to diagnose all undiagnosed men in this sub-population in the absence of new infections. In contrast, in the overseas born male-to-male sexual contact group the TDF, YDF, and CDR all stagnated over the same period. Amongst people living with HIV who reported heterosexual contact, the TDF has increased for both women and men, but remain below the global target of 95%. For women, the CDR is high, at 1.76, meaningly almost two women being diagnosed for every new infection in 2019. Despite little change in the number diagnosed each year, new infections have fallen and a higher proportion of women who are undiagnosed each year received a diagnosis (YDF). In contrast, for heterosexual men, the CDR is below one meaning new infections exceeded diagnoses. Even though the TDF and YDF increased for heterosexual men, the CDR falling below one implies a growing undiagnosed population.

The YDF and CDR are useful as a complimentary measure to the TDF. Although the trends tend to align with the TDF, changes of public health significance resulting from prevention and testing strategies will be more clearly shown in the TDF and CDR because they focus on changes during the previous year rather than all people living with HIV (who may have been infected a long time ago). Divergences between sub-populations also become apparent earlier. In our study, changes in the TDF and CDR were most observable among the Australian-born men who reported male-to-male sexual contact, coinciding with increased HIV testing programs targeted towards gay, bisexual, and other men who have sex with men over the last decade and the scale-up of PrEP since 2016.[21,22]

The differences observed across the sub-populations highlight the need for an ongoing focus on HIV testing strategies, enhanced efforts to decrease the time between acquisition and diagnosis, and to ensure equity among sub-populations who are testing infrequently.[23–25] The falling TDF and CDR for overseas born men reporting male-to-male sexual contact is particularly concerning, indicating the number of undiagnosed people and the time required to diagnose all undiagnosed people in this sub-population is increasing.

Our study is subject to certain limitations. Firstly, YDF and CDR estimates are susceptible to fluctuations and high uncertainty, especially in the most recent years and for populations with small numbers of diagnoses. The CDR also relies on an accurate estimation of new infections that occur within a country excluding those who acquire HIV prior to arrival. Secondly, when using the ECDC HIV modelling tool to infer new infections, we excluded diagnoses among people who had previously been diagnosed overseas but were unable to exclude diagnoses where HIV was acquired overseas but diagnosis first occurred in Australia, which may make up to 34% of diagnoses among people born overseas.[18] This means our estimates for new infections will likely be an overestimate and the CDR will be underestimated. Thirdly, our study only considered four key sub-populations (Australian-born and overseas-born individuals having reported male-to-male sexual contact, and heterosexual men and women). It is crucial to consider all sub-population for more accurate predictions and modifications to HIV prevention campaigns nationally, however this also needs to consider the uncertainty associated with small numbers of diagnoses. Because this is the first study utilizing the YDF and CDR in Australia, conducting a similar analysis on a more routine basis (e.g., every five years) would strengthen the use of the estimates and ease of interpretation for national surveillance reporting.

To our knowledge, our study is one of the first to use the CDR for HIV and apply the YDF to key populations. While the TDF has been widely used for HIV, as far as we are aware the YDF has only been used in a limited number of studies for HIV [8,10] despite a call for its application to support measuring global HIV targets.[6] The CDR has been integrated into targets for global tuberculosis control and has been applied in numerous settings.[26–30] The only other infectious disease we are aware of using the CDR is leprosy.[31,32] Its significance for understanding epidemics and predicting outcomes has also been highlighted due to the COVID-19 pandemic.[33] The addition of estimates for YDF and CDR to Australian HIV surveillance will be valuable because Australia’s HIV epidemic is at a point where up-to-date data for recent periods are required to determine progress towards the goal of elimination of HIV transmission. Routine use of these metrics would enhance our understanding of the HIV response, at a critical juncture, where we are focusing on achieving the goal of elimination of HIV transmission.

## Conclusions

Our study estimated trends in undiagnosed HIV in Australia over a 12-year period, 2008-2019. It shows additional analyses, and measures, such as the YDF and the CDR, can help to build a more comprehensive and accurate picture of the impact of HIV interventions on specific sub-populations. Incorporating these new metrics into HIV surveillance will help to further evaluate the effectiveness of testing campaigns and other HIV focused public health messaging, and to ensure efforts are appropriately targeted to reduce new infections in key populations.

## Data Availability

The code used to produce the estimates with aggregate data and results for this study are available online at https://zenodo.org/record/7058987. Updated code available from https://github.com/The-Kirby-Institute/Cascade_calculations.

https://github.com/The-Kirby-Institute/Cascade_calculations.

## Authors’ contributions

RG, HC, NM conceptualized the study and data analysis plan. HC, LK, SM, RTG extracted and collected data. RTG wrote the cascade and metric calculation code, produced the metric estimates and plots, and manages the online code repository. LK performed the statistical analyses. HC and RTG drafted the manuscript. AG, RG, SM and NM reviewed and provided input into manuscript drafts. All authors read, commented on, and approved the final manuscript.

## Acknowledgments

The authors thank Shawn Clackett for earlier investigations into the use of the YDF and CDR metrics and Dr Hamish McManus for statistical advice. The authors acknowledge the contribution of the members of the Australian HIV Diagnosis and Care Cascade reference group and acknowledge people living with HIV and their invaluable contribution to research.

## List of abbreviations

CDR: Case detection rate
TDF: Total diagnosed fraction
YDF: Yearly diagnosed fraction

